# Identification of a *de novo* mutation in *TLK1* associated with a neurodevelopmental disorder and immunodeficiency

**DOI:** 10.1101/2023.08.22.23294267

**Authors:** Marina Villamor-Payà, María Sanchiz-Calvo, Jordann Smak, Lynn Pais, Malika Sud, Uma Shankavaram, Alysia Kern Lovgren, Christina Austin-Tse, Vijay S Ganesh, Marina Gay, Marta Vilaseca, Gianluca Arauz-Garofalo, Lluís Palenzuela, Grace VanNoy, Anne O’Donnell-Luria, Travis H. Stracker

## Abstract

**Background:** The Tousled-like kinases 1 and 2 (TLK1/TLK2) regulate DNA replication, repair and chromatin maintenance. TLK2 variants are associated with ‘Intellectual Disability, Autosomal Dominant 57’ (MRD57), a neurodevelopmental disorder (NDD) characterized by intellectual disability (ID), autism spectrum disorder (ASD) and microcephaly. Several TLK1 variants have been reported in NDDs but their functional significance is unknown.

**Methods:** A male patient presenting with ID, seizures, global developmental delay, hypothyroidism, and primary immunodeficiency was determined to have a novel, heterozygous variant in TLK1 (c.1435C>G, p.Q479E) by genome sequencing (GS). Single cell gel electrophoresis, western blot, flow cytometry and RNA-seq were performed in patient-derived lymphoblast cell lines. In silico, biochemical and proteomic analysis were used to determine the functional impact of the p.Q479E variant and previously reported NDD-associated TLK1 variant, p.M566T.

**Results:** Transcriptome sequencing in patient-derived cells confirmed expression of TLK1 transcripts carrying the p.Q479E variant and revealed alterations in genes involved in class switch recombination and cytokine signaling. Cells expressing the p.Q479E variant exhibited reduced cytokine responses and higher levels of spontaneous DNA damage but not increased sensitivity to radiation or DNA repair defects. The p.Q479E and p.M566T variants impaired kinase activity but did not strongly alter localization or proximal protein interactions.

**Conclusion:** Our study provides the first functional characterization of TLK1 variants associated with NDDs and suggests potential involvement in central nervous system and immune system development. Our results indicate that, like TLK2 variants, TLK1 variants may impact development in multiple tissues and should be considered in the diagnosis of rare NDDs.

## Introduction

The Tousled-like kinases 1 and 2 (TLK1 and TLK2) are conserved serine-threonine kinases that function in numerous cellular processes, including DNA replication, DNA repair, transcription and chromatin maintenance(1). Both TLK1 and TLK2 interact with and regulate the ASF1A and ASF1B histone H3-H4 chaperones(2–7). Depletion of both TLK1 and TLK2, or both ASF1A and ASF1B, led to overlapping cellular phenotypes, including DNA damage, innate immune activation and the induction of the alternative lengthening of telomeres (ALT) pathway(8–10). TLK1 and TLK2 are regulated by the DNA damage response and the phosphorylation of TLK1 on its C-terminus by the checkpoint kinase CHK1 inhibits TLK1 activity(11,12). TLK1 also phosphorylates the RAD9 protein, part of the RAD9-RAD1-HUS1 (9-1-1) complex that responds to DNA damage and regulates CHK1 activity, as well as the NEK1 kinase, that is involved in the DNA damage response and ciliogenesis(4,13–19).

Despite clear associations with fundamental cellular processes and multiple links to the DNA damage response, *Tlk1* deficiency in mice did not result in any overt phenotypes. Notably, both immune system development and fertility were grossly normal, suggesting DNA repair-dependent processes in V(D)J recombination and meiosis were functional(8). Additionally, the deletion of *TLK1* in hundreds of cancer cell lines did not strongly influence cell fitness(20). In contrast, loss of *Tlk2* in mice led to embryonic lethality due to a role in placental development, and *TLK2* is an commonly essential gene in many cancer cell lines, indicating distinct functions or regulation that remain to be fully understood(8,20).

Variants in *TLK2* have been implicated in Intellectual Developmental Disorder, Autosomal Dominant 57 (MRD57, MIM# 618050), a heterogenous neurodevelopmental disorder (NDD) characterized by intellectual disability (ID), autism spectrum disorder (ASD), microcephaly, additional behavioral problems, growth delay and facial dysmorphism, including blepharophimosis, telecanthus, prominent nasal bridge, broad nasal tip, thin vermilion of the upper lip and upslanting palpebral fissures(21–25). A subset of cases also exhibited gastrointestinal problems, seizures, skeletal malformations and ocular problems. With one exception, all of the disease-associated *TLK2* variants identified to date are heterozygous, and those examined biochemically showed diminished kinase activity, suggesting pathological outcomes are largely due to reduced kinase activity(23,24).

Activation of TLK2 requires dimerization through its first coiled-coil (CC1), which is also required for its heterodimerization with TLK1(26,27). As TLK1 and TLK2 interact, some of the identified *TLK2* missense variants may exert dominant negative effects on wild type TLK2 and potentially TLK1(28). While >40 *TLK2* variants have been identified in NDD patients, to date, only four *de novo TLK1* variants have been associated with NDDs (Table 1)(21–25,29). However, no causal links have been established, and the effects of any of the individual variants on protein function have not been investigated.

**Table 1:** List of *TLK1* variants reported with links to human disease. Positions are based on <u>NM_012290.5</u>.

| *TLK1* variant | TLK1 protein | Disease association | Reference |
| --- | --- | --- | --- |
| c.74C>T | p.P25L | Intellectual disability | (25) |
| c.112_113del | p.T38fs | Intellectual disability | ClinVar |
| c.424G>A | p.R142S | Limb, ear, eye and muscle abnormalities | DECIPHER |
| c.1101del | p.K367Nfs* | Schizophrenia | (30) |
| c.1435C>G | p.Q479E | Intellectual disability, microcephaly, immunodeficiency | This study |
| c.1697T>C | p.M566T | Autism Spectrum Disorder | (29) |
| c.1796C>G | p.A599G | Congenital heart defect | (31) |

Here we report the identification of a male proband who presented with microcephaly, ID, seizures, global developmental delay, cerebral calcifications, feeding difficulties, growth hormone deficiency, hypothyroidism, urticaria and primary immunodeficiency. He had extensive prior genetic testing, including whole exome sequencing (WES), that did not identify a clear genetic cause for his phenotype. Subsequent research exome sequencing (ES) and genome sequencing (GS) through the Rare Genomes Project (RGP) identified heterozygous, *de novo* variants in *TLK1* c.1435C>G (p.Q479E) and *MDM1* c.1197dupT (p.K400Ter).

## Materials and Methods

### Genome sequencing (GS), variant calling, and prioritization

GS and data processing for this individual and his parents were performed by the Genomics Platform at the Broad Institute of MIT and Harvard. PCR-free preparation of sample DNA (350 ng input at >2 ng/uL) was accomplished using Illumina HiSeq X Ten v2 chemistry. Libraries were sequenced to a mean target coverage of >30X. WGS data was processed through a pipeline based on Picard, using base quality score recalibration and local realignment at known indels. The BWA aligner was used for mapping reads to the human genome build 38. Single Nucleotide Variants (SNVs) and insertions/deletions (indels) were jointly called across all samples using the Genome Analysis Toolkit (GATK) HaplotypeCaller package version 4.0. Default filters were applied to SNV and indel calls using the GATK Variant Quality Score Recalibration (VQSR) approach. Annotation was performed using Variant Effect Predictor (VEP). GATK-SV(32) was used to detect structural variants (SVs), which were annotated with the GATK SVAnnotate tool. Mitochondrial DNA (mtDNA) single nucleotide and small indel variants were called from GS data using the gnomAD-mitochondria pipeline(33) and large mtDNA deletions were called by MitoSAlt(34). ExpansionHunter v5 was used to genotype known disease-associated short tandem repeats (STRs)(35). Lastly, the variant call set was uploaded to seqr for analysis by the RGP team. Variants in *TLK1* (p.Gln479Glu) and *MDM1* (p.Lys400Ter) were submitted to ClinVar (submission ID: SUB13580789).

### Cell culture and generation of patient cell lines

Peripheral blood mononuclear cells were isolated from whole blood using Histopaque-1077 (Sigma-Aldrich) and subsequently immortalized with Epstein-Barr virus transformation (Coriell Institute). Lymphoblastoid cell lines were cultured in RPMI-1640 medium (Corning) supplemented with 15% fetal bovine serum (FBS) (Gibco), 1% penicillin-streptomycin, and 2 mM L-glutamine. AD-293 cells (Stratagene) were grown in DMEM supplemented with 10% FBS (Sigma-Aldrich), 50□U/mL penicillin and 50□µg/mL streptomycin (Thermo Fisher Scientific) and authenticated using STR testing (ATCC). All cells were kept at 37°C in a 5% CO_2_ incubator. Cells were counted with a Countess cell counter (Invitrogen) and viability was assessed using trypan blue. For any given experiment, only cell cultures with a viability >90% were used. Cells were routinely tested for mycoplasma and found negative (Lonza).

### Genomic DNA and RNA Purification

Genomic DNA was purified from **lymphoblastoid cell lines** using the Gentra Puregene Cell kit (Qiagen) following manufacturer’s instructions. Total RNA was extracted from lymphoblastoid cell lines (LCLs) using the SV Total RNA Isolation System (Promega) following manufacturer’s instructions. Full methods provided in **online supplemental materials and methods**.

### PCR amplification and DNA extraction from agarose gels

Genomic DNA was subjected to 35 cycles of PCR amplification using Q5 High-Fidelity DNA Polymerase (New England Biolabs) with primers that allow for the amplification of the region containing the variants of interest (**online supplemental table S1).** PCR products were run in a 2% agarose gel and purified using the Nucleospin Gel and PCR clean-up kit (Macherey-Nagel) following manufacturer’s instructions. Full methods provided in **online supplemental materials and methods.**

### Nextera library preparation and sequence analysis for variant calling in PCR products

DNA libraries were prepared using the Nextera XT DNA Library Prep kit (Illumina) following manufacturer’s instructions. Sequencing of PCR amplicons was carried out using the MiSeq Nano kit sequencing system (Illumina) with 150 bp paired-end (PE) reads. Cleaned reads were obtained by Trim Galore and FASTQ sequences aligned to the hg38 genome build followed by BCFtools for variant calling and alternate allele frequency of the sequenced genes(36). A filter with quality score >=20 was used to remove variants likely observed purely by chance.

### RNA sequencing (RNA-seq)

RNA-seq was performed in the CCR Genomics core facility. RNA-seq data processing was performed using the RNA-seq pipelines of the CCBR Pipeliner framework (https://github.com/CCBR/Pipeliner). Full analysis methods provided in **online supplemental materials and methods.** Data was submitted to GEO: GSE241032.

### *In silico* prediction and modeling of missense variants

*In silico* prediction of variants was analyzed using seqr (https://seqr.broadinstitute.org)(37). Residue conservation was determined using ConSurf (https://consurf.tau.ac.il)(38) and the heatmap generated using Prism. The TLK1 kinase domain structure was generated using Alphafold (https://alphafold.ebi.ac.uk) and color modified in PyMol to indicate residues of interest.

### Comet and cell proliferation assays

Comet assays were performed according to manufacturer’s instructions (Trevigen). Proliferation was assessed using the Click-iT Plus EdU Flow Cytometry Assay Kit (Invitrogen) following manufacturer’s instructions. Full methods provided in **online supplementary materials and methods.**

### Cloning, site-directed mutagenesis, transfection, streptavidin affinity purification, western blotting and kinase assays

Cloning, mutagenesis, transfections, western blotting, streptavidin affinity purification of TLK1 from cell lysates and *in vitro* kinase assays were performed as previously described with minor variations(24). Full methods are provided in the online supplemental materials and methods. Primers are provided in **online supplementary table S1,** antibodies are provided in **online supplementary table S2** and uncropped westerns including those used for quantification are shown in **online supplementary figure S1.** Westerns and gels used for kinase assays are shown in **online supplementary figure S2.** Proteomic data is available in the PRIDE repository: <u>PXD019450</u>.

## Results

### Identification of a proband with a neurodevelopmental disorder and a *de novo TLK1* variant

After an uncomplicated pregnancy, the proband was born with microcephaly noted early in the postnatal period. Between 0-5 months of age, he was admitted to the neonatal intensive care unit (NICU) due to an infection that progressed to respiratory failure. During a several month course in the NICU, he was found to have B-and T-cell deficiency and diagnosed with primary hypothyroidism (TSH 33 uIU/mL, repeat 18.6 uIU/mL). At less than 5 months old, he had a stroke and a generalized seizure and was treated with levetiracetam, after which he was weaned off levetiracetam. Brain MRI revealed symmetric periventricular and subcortical leukoencephalopathy with mild cystic white matter encephalomalacia, ex vacuo ventriculomegaly, thinning of the corpus callosum, and bilateral optic nerve atrophy (**Figure 1A-D**.

**Figure 1:**
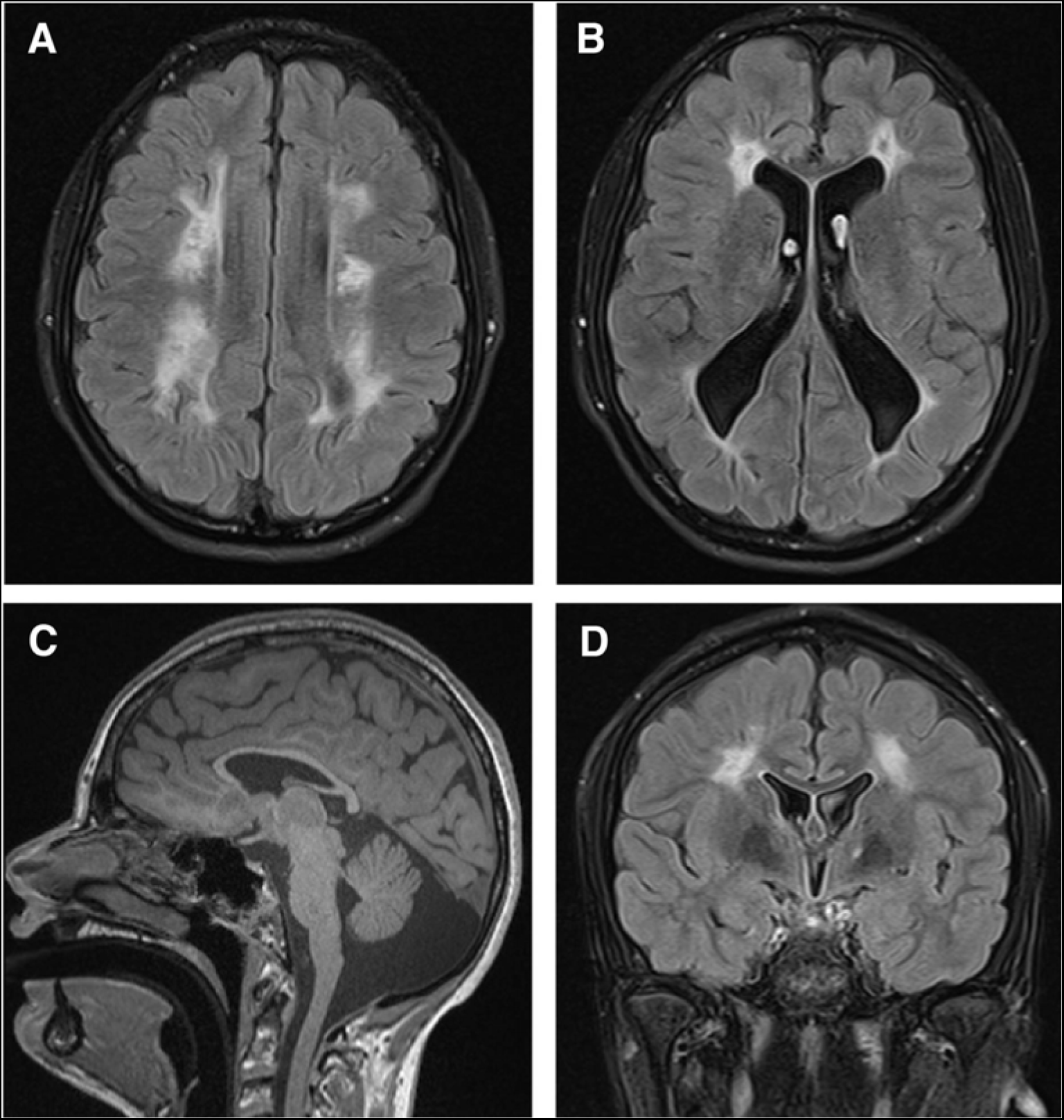
T2/FLAIR axial (A, B), sagittal (C), and coronal (D) sections from age 10-15 years shows symmetric periventricular and subcortical leukoencephalopathy with mild cystic white matter encephalomalacia, ex vacuo ventriculomegaly, and thin corpus callosum.

T cells normalized before 5 years of age, but he still has B-cell deficiency and hypogammaglobulinemia (low IgG), treated with weekly subcutaneous immunoglobulin. Enzymatic testing for severe combined immunodeficiency (SCID), including adenosine deaminase (ADA) and purine nucleoside phosphorylase (PNP), was normal (age 0).

Between 15 and 20 months, he had a kidney biopsy following episodes of proteinuria and hematuria that was not diagnostic. Between 6 and 10 years old, he was diagnosed with growth hormone deficiency and treated with growth hormone injections.

His medical history is also notable for significant global developmental delay, hypotonia, mild outer retinal dystrophy, feeding difficulties, and skin findings including poikiloderma, annular urticaria, and a chronic rash that flares up upon infection. He has used a walker for support from 15-20 months of age and has significant speech and language delays, though his receptive abilities are more developed than his expressive language.

The proband had thorough clinical genetic testing, all of which came back normal. This included chromosomal microarray and karyotyping, candidate-gene sequencing (*CIAS1, RAB27A, RECQL4*), targeted mutation analysis (Factor V Leiden p.R506Q, Factor II/prothrombin p.G20210A), mtDNA analysis and nuclear mitochondrial disease panel, and panel testing (SCID panel, Aicardi Gouitéres panel, Noonan syndrome panel, dyskeratosis congenita panel). Clinical exome sequencing (ES) and subsequent ES reanalysis in 2017 were negative.

Research GS done through RGP identified a rare *de novo* frameshift variant in *MDM1* (c.1197dupT; p.Lys400Ter; 12:68315249, hg38) and a rare *de novo* missense variant in *TLK1* (c.1435C>G; p.Gln479Glu; 2:171007045, hg38) further detailed in this report.

### Characterization of patient-derived cells expressing the *TLK1* and *MDM1* variants

To further investigate the potential impact of the mutations, PBMCs from the proband and an unaffected female sibling of a similar age, were isolated and transformed with Epstein Barr Virus (EBV) to generate lymphoblastoid cell lines (LCLs). Genomic DNA from expanded LCLs was amplified with specific primers, and sequencing confirmed the presence of both the *TLK1* and *MDM1* variants identified by GS.

We next performed RNA-seq on each LCL to identify differentially expressed genes (DEGs) and assess the expression levels of the specific variant alleles of both *MDM1* and *TLK1*. Both *TLK1* and *MDM1* were expressed to similar levels in each cell line (**Figure 2A and online supplementary table S3**). The allele harboring the *TLK1* variant was transcribed to a similar extent as the wildtype allele (**Figure 2B** and **online supplementary table S4**), and quantitative western blotting showed total TLK1 protein levels slightly elevated in cells from the proband with the p.Q479E variant compared to the unaffected sibling (**Figure 2C**). In contrast to the *TLK1* variant, the *MDM1* mutant allele was not detectably expressed, likely indicating nonsense-mediated RNA decay (NMD) (**Figure 2B**). However, total MDM1 protein levels appeared similar, consistent with RNA-seq data, suggesting that MDM1 protein levels are unaffected in the LCL line (**Figure 2D**). We next examined the phosphorylation of two reported TLK1 substrates ASF1A and NEK1(6,17). In both cases, no significant difference in phosphorylation of either substrate could be observed between the cell lines (**Figure 2E-F**).

**Figure 2:**
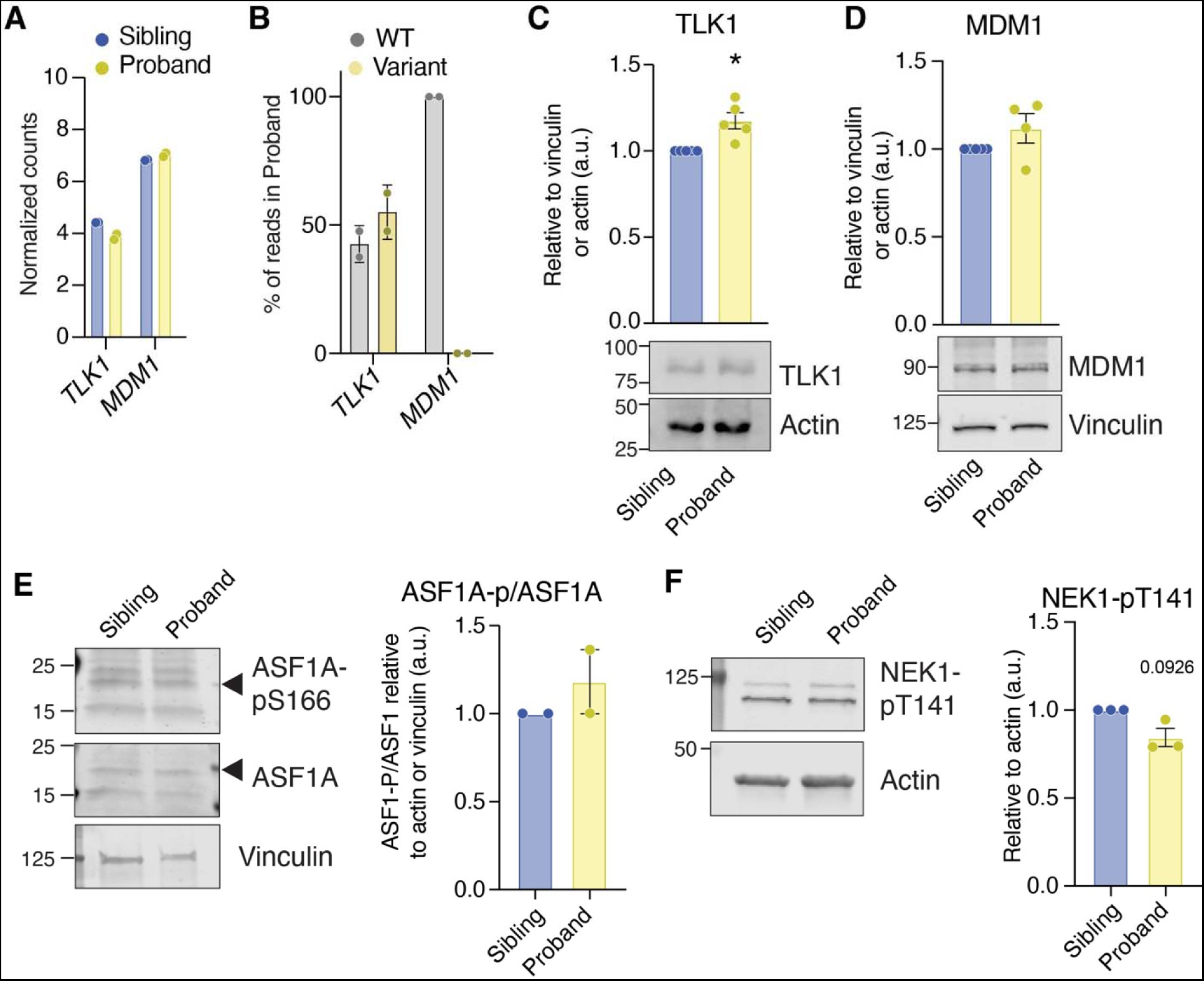
*TLK1* and *MDM1* variant alleles and impact on protein levels. **A.** Normalized read counts for the indicated genes from RNA-seq of transformed LCLs of the Sibling or Proband. N=2, mean and standard deviation are shown. Full details in **online supplementary table S3**. **B.** Allele specific expression of the indicated genes in the Proband inferred from RNA-seq data. N=2, mean and standard deviation are shown. Full details in **online supplementary table S4. C.** Representative blot of protein levels of TLK1. Quantification of western blots (N=5) is shown in right panel normalized to Vinculin or Actin. **D.** Representative blot of protein levels of MDM1 and quantification of western blots (N=4) is shown in right panel normalized to Vinculin or Actin. **E.** Representative blot of ASF1A and ASF1A-pS166 (ASF1A-p) levels and quantification of western blots (N=2) is shown in right panel. **F.** Representative blot NEK1-pS141 (P-NEK1) levels and quantification of western blots (N=3) is shown in right panel. N represents biological replicates, statistical significance was determined using an unpaired t test with Welch’s correction (*P<0.05) in panels C,D.

### Gene expression differences consistent with immunodeficiency

RNA-seq data was further analyzed to identify DEGs between the LCL cell lines (**Figure 3A**). Ingenuity pathway analysis (IPA) identified Primary Immunodeficiency Signaling (PID) as a top enriched category, consistent with the immunodeficiency of the proband (**Figure 3A-C**). Many of the genes identified are involved in class switch recombination (CSR), and additional DEGs in CSR genes were identified manually, including CD27 and FCRL3 (**Online supplemental table S3**). In addition, the STAT3 pathway, Osteoarthritis pathway (OSTEO) and Hepatic fibrosis pathways were among the top 4 significantly altered pathways in the LCLs of the proband compared to the sibling (**Figure 3A-C**). Notably, these pathways included a number of transcriptional regulators, cytokines and other cell surface proteins that influence cell to cell interactions in the immune system.

**Figure 3:**
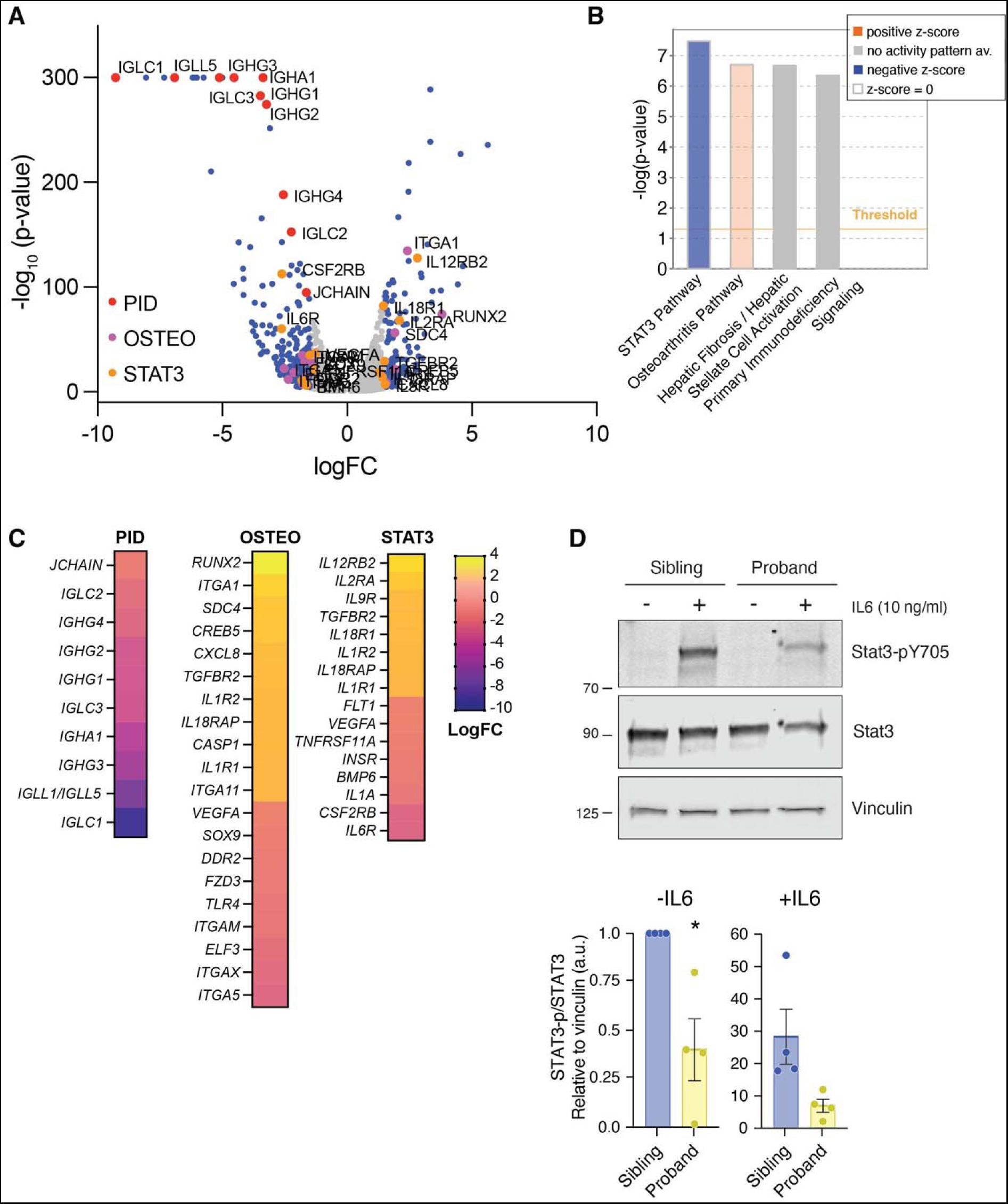
Gene expression differences identify defects in STAT3 signaling in patient-derived cell lines. **A.** Volcano plot of RNA-seq data depicting DEGs in the Proband compared to Sibling LCLs. Duplicate samples from each LCL line were analyzed. Full data in **online supplemental Table S3**. Genes in each enriched category identified in IPA analysis (B) are shown in the indicated color. Hepatic fibrosis genes overlap with Osteoarthritis and are therefore not shown. **B.** Ingenuity Pathway Analysis (IPA) of RNA-seq data is shown. **C.** Heatmap of individual genes from the indicated enriched pathway are shown. **D.** Analysis of STAT3 phosphorylation on Y705 in response to IL6 in LCLs. Quantification of 4 independent experiments is shown below the western blot. STAT3-p (Y705)/STAT3 was normalized to vinculin and samples were normalized to Sibling -IL6. Statistical significance was determined using an unpaired t test with Welch’s correction (*P<0.05).

Examination of the STAT3 signaling related genes revealed a number of DEGs, including the Interleukin-6 receptor (IL6R) and CSF2RB, which is a component of multiple cytokine receptors. As mutations in STAT3 and IL6R are associated with distinct immunodeficiency syndromes (39), we analyzed STAT3 signaling in cells treated with IL6 and examined the phosphorylation of Tyrosine 705 (Y705) of STAT3, a marker of its activation. While increased levels of pY705- STAT3 were observed in both samples following IL6 treatment, it was markedly reduced in the proband, indicating reduced sensitivity to IL6 (**Figure 3D**). Together, these data indicated that the cells from the proband showed gene expression changes consistent with immunodeficiency and defective CSR and were less responsive to IL6 cytokine stimulation.

### Growth defects and increased DNA damage in a patient-derived cell line

To determine the potential impact of the genetic variants and DEGs on cell growth and viability, we compared the growth of the proband-and sibling-derived cell lines by plating the same number of live cells and counting them every 24 hours over 72 hours. Viability at the beginning of the experiment was always >90%. The proband-derived cells showed reduced growth over 72 hours, with observable differences after 48 hours (**Figure 4A**). Flow cytometry analysis of EdU-pulsed cultures showed a reduction of cells in S-phase with a compensatory increase in the G1 population (**Figure 4B and 4C**). Both the G2/M and sub-G1 populations were also increased in cells from the proband (Figure 4B and 4C). As this suggested potential activation of cell cycle checkpoints, we examined p53 and p21 levels and found that both were upregulated in the proband compared to the unaffected sibling (**Figure 4D and 4E**).

**Figure 4:**
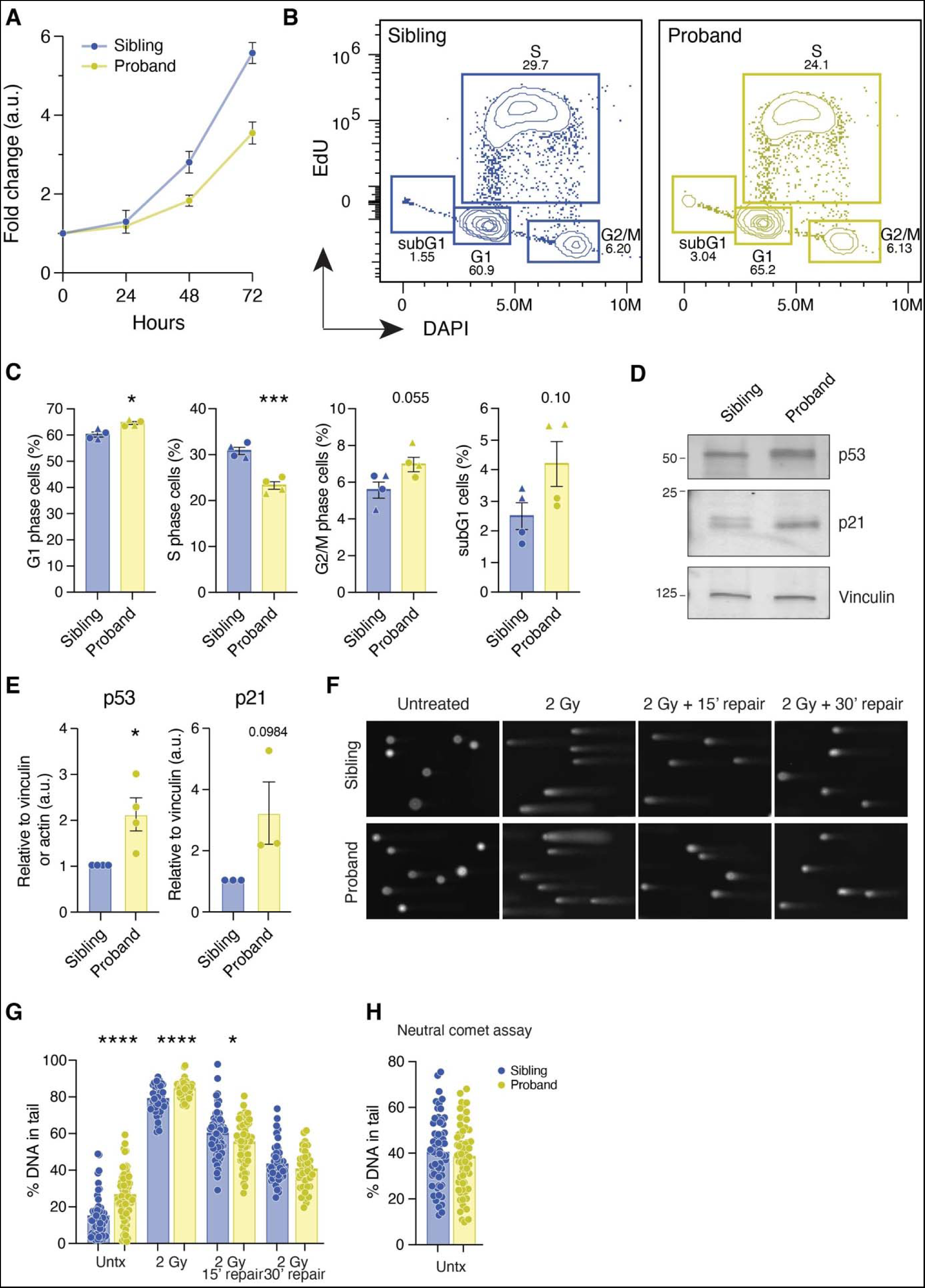
Analysis of cell cycle and DNA damage in patient cell lines. **A.** Relative cell growth of the Proband and Sibling LCLs. N=6. **B.** Example of representative flow cytometry data to analyze cell cycle in LCLs. Cells were pulsed with 10 µM EdU for 1 hour and stained with DAPI for DNA content. **C.** Quantification of cell cycle phases from N=2 independent experiments with 2 biological replicates each. Replicates from same experiment indicated with a triangle or circle. **D.** Western blot analysis of p53 and p21 levels in LCLs. **E.** Quantification of p53 (N=4) and p21 (N=3) levels from biological replicates. **F.** Representative images of alkaline comet assays untreated or treated with the indicated dose of ionizing radiation (IR). **G.** Analysis of tail moment with the indicated treatments and recovery times. Representative of N=2 independent experiments. **H.** Analysis of tail moment in neutral comet assays. Representative of N=2 independent experiments. At least 100 comets were analyzed per condition and experiment. Statistical significance was determined using an unpaired t test with Welch’s correction (****P<0.0001, ***P<0.001, **P<0.01, *P<0.05).

As the increased p53/p21 levels and elevated sub-G1 population suggested stress or DNA damage signaling, we used the alkaline comet assay to examine the LCLs for spontaneous DNA damage and to examine DNA repair following exposure to 2 Gy ionizing radiation (IR). The proband-derived cells showed increased tail moment values in the alkaline comet assay in the absence of IR treatment, indicating elevated levels of DNA damage (**Figure 4F and 4G**). However, examination using the neutral comet assay did not reveal evidence of increased DNA strand breaks (**Figure 4H**). Consistent with an absence in DSB repair defects, comets induced by IR treatment were resolved similarly to those in the control cell line, indicating that overall DSB repair was functional, suggesting that spontaneous damage may be ssDNA gaps or base damage (**Figure 4G**).

### *In silico* and biochemical analysis of the *TLK1* variant

As the *TLK1* variant was the best genetic candidate for the observed phenotypes, we analyzed it with a variety of prediction tools to determine if the variant was potentially damaging to TLK1 activity (37). The results were variable, with some prediction tools indicating that the variant was damaging and others indicating it was tolerated (**online supplementary table S5).** Analysis of missense constraint (www.decipher.com) indicated that the kinase domain of TLK1 is highly constrained (**online supplementary figure S3**). We next analyzed the specific conservation of the Q479 residue using Consurf, including also an additional *TLK1* variant that was previously reported in a patient with ASD, p.M566T (Table 1)(29,38). Both residues are located within the kinase domain of TLK1, with Q479 located in a beta sheet of the N-lobe and M566 in an alpha helix of the C-lobe. Both residues scored as highly conserved, indicating that they could be important for kinase activity (**Figure 5A**).

**Figure 5:**
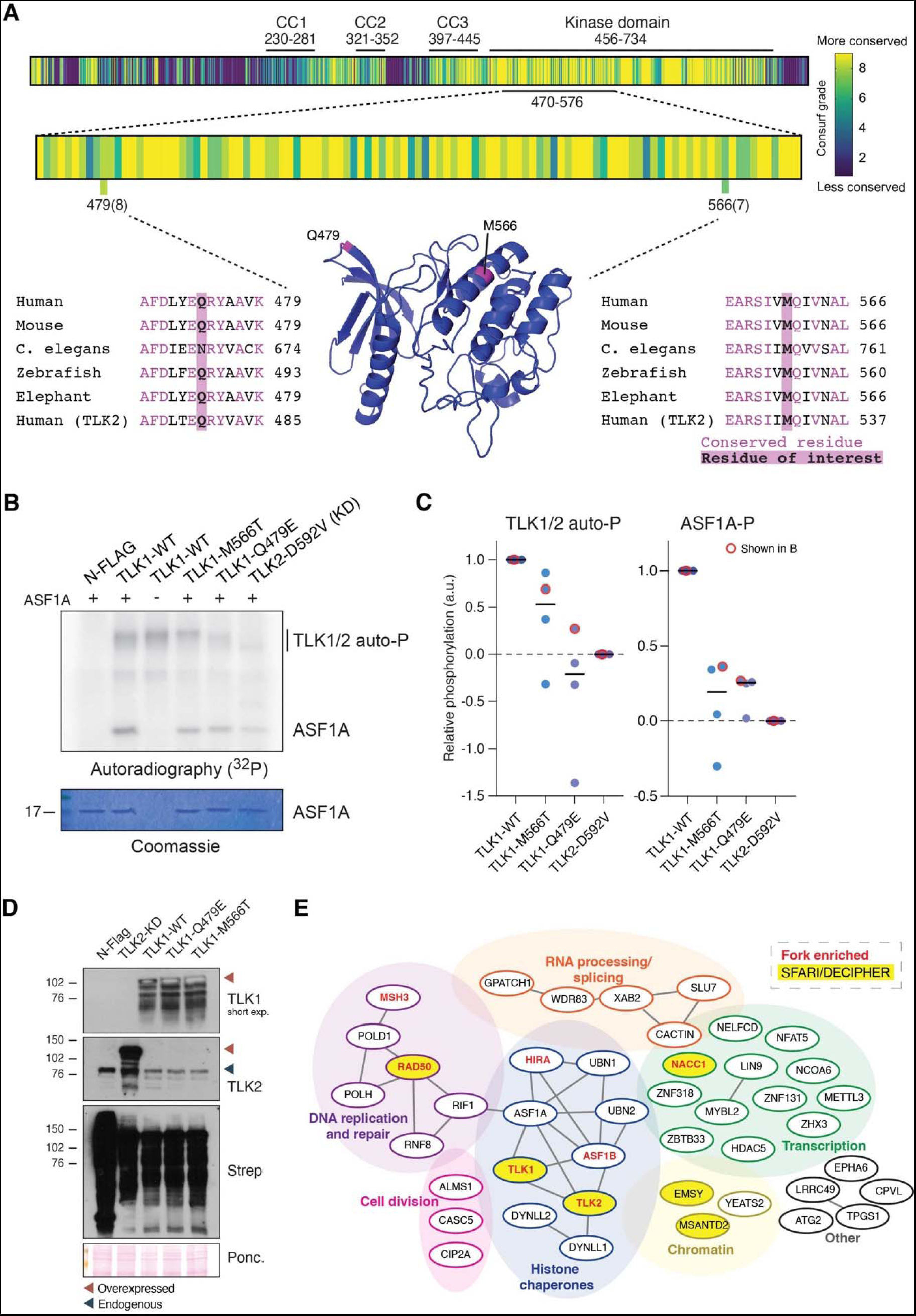
Conservation analysis, kinase activity and proximal interactions of NDD associated TLK1 variants. **A.** Consurf was used to analyze the level of conservation of the amino acids of human TLK1, with both Q479 and M566 scoring as highly conserved. The predicted structure of the TLK1 kinase domain (Alphafold) is shown with the location of the two residues highlighted in pink(42,43). **B.** Representative *in vitro* kinase assay of Streptavidin purified TLK1-WT or NDD variants on purified ASF1A. TLK2-KD is used as a negative control. **C.** Quantification of N=4 independent kinase assay experiments. **D.** Western blotting of transfected BioID constructs and biotin labeling imaged with Streptavidin. Ponceau is provided as a transfer control. **E.** Network depiction of the proximal interactors identified with TLK1 with physical interactions indicated by solid lines. Proteins identified on nascent DNA at replication forks by iPOND-MS and proteins in the Simon’s Foundation Autism Research Initiative (SFARI) or DECIPHER databases are indicated(44). Full results provided in **online supplementary table S6** and additional data in **online supplementary figure S4.**

To directly determine if kinase activity was affected, the p.Q479E or p.M566T variants were generated in an expression vector using site-directed mutagenesis. N-terminally Strep-FLAG-tagged TLK1-WT, or the two variants, were expressed in AD-293 cells and affinity purified from cell lysates for *in vitro* kinase assays using purified ASF1A as a substrate (**Figure 5B**). A kinase dead mutant of TLK2 (D592V) was used as a negative control for non-specific labeling of the substrate(24). Both the p.Q479E and p.M566T variant proteins showed reduced autophosphorylation activity compared to TLK1-WT and their kinase activity was impaired to similar extent as TLK2-KD compared to TLK1-WT (**Figures 5B-C**). These results indicated that both variants resulted in severely impaired TLK1 kinase activity.

### The NDD-associated TLK1 variants do not strongly alter the proximal proteome

We next examined the proximal proteomes of the TLK1 variants using BioID-MS. The BioID enzyme was fused to the N-terminus of the wild type (WT), p.Q479E and p.M566T constructs, and they were expressed by transient transfection in AD293 cells that were pulsed with biotin for 24 hours (**Figure 5D**)(40). Cells were harvested and biotinylated proteins purified using Streptavidin beads. Isolated proteins were subjected to mass spectrometry and analyzed using SAINTexpress to identify proteins specifically labeled by TLK1-WT or either variant allele (**online supplementary table S6**)(41). The overall network of proximal interactors identified was similar to that we previously observed for TLK2 (**Figure 5E and online supplementary figure S4**)(24). Few strong differences in proximal interactors with either p.Q479E and p.M566T were identified, including with substrates ASF1A and ASF1B, or DYNLL1/LC8, which we previously identified as a robust interactor of TLK2 (**online supplementary figure S4 and table S6**) (8) (24). These results indicated that while both TLK1 variants had reduced activity, their proximal interactions were not strongly altered, in contrast to what was observed with several TLK2 NDD variants we previously reported(24).

## Discussion

In this report we describe a proband with a complex phenotype including developmental delay, intellectual disability, leukoencephalopathy, and primary immunodeficiency. Genetic testing identified potential candidate genes, including *TLK1* and *MDM1*. The *MDM1* variant was not detectable in LCLs, but total *MDM1* expression levels did not appear to be strongly altered at the mRNA or protein level (**Figure 2**). However, we cannot rule out a potential impact of *MDM1* haploinsufficiency in other tissues. In mice, a homozygous truncation mutation of *Mdm1* was linked to retinal degeneration(45). Therefore, the optic nerve atrophy or retinal dystrophy observed in the proband could reflect defects in MDM1 function due to heterozygous expression of the truncated allele or haploinsufficiency in eye development. However, we suggest that *TLK1* is the most likely genetic contributor to the overall phenotype, particularly the neurodevelopmental outcomes.

While TLK1-deficient mice do not exhibit any obvious developmental phenotypes, ample evidence exists that kinase dead forms of TLK1 may impact the function of TLK1 or TLK2 through heterodimerization via the CC1 motif, resulting in a dominant negative effect(26–28). We examined several substrates of TLK1, ASF1A and NEK1, and did not find a clear difference in their phosphorylation in patient-derived cell lines (**Figure 2**). However, cells exhibited significant levels of spontaneous DNA damage and growth defects (**Figure 4**), indicating potential effects on other substrates that we did not analyze in this study. Recent work identified RAD54, a protein involved in homologous recombination of DSBs, as a TLK1 substrate(46). However, the increased DNA damage we observed was not DSBs. TLK1 BioID identified two proteins involved in Mismatch Repair (MMR), MSH3 and POLD1, as proximal interactors (**Figure 5**). The MMR pathway is involved in the repair of some types of base damage, including that caused by elevated reactive oxygen species, and is also required for CSR(47,48). Thus, TLK1 could potentially play an undefined regulatory role in MMR that is compromised by the variant allele.

Immunodeficiency has not been reported in mouse models of TLK1 or TLK2 deficiency and is therefore hard to directly link to the heterozygous *TLK1* variant identified in this patient based on current knowledge(8). The immunological presentation, as well as the RNA-seq data generated from PBMCs and LCLs, is consistent with defects in lymphocyte maturation, particularly in B-cells, reflecting the proband’s status (**Figure 3**). While the proband initially presented with primary immunodeficiency, T-cells recovered, indicating that remaining issues may be the result of impaired B-cell maturation. CSR is a B-cell specific process that is dependent on a number of non-homologous end-joining (NHEJ) pathway genes that largely, but do not completely, overlap with those required for V(D)J recombination(49). A particular feature of CSR is a distinct requirement for 53BP1 and RIF1 that work with a number of proteins to prevent resection and enforce NHEJ(50). Recent work directly linked ASF1 to RIF1-dependent NHEJ DNA repair(51–53). We, and others, have identified RIF1 as a proximal interactor with TLK1 and TLK2 (8,52–54). Thus, defects in TLK1 that influence ASF1 function in this pathway could potentially impact CSR. Further work remains to determine how important TLK-mediated regulation of ASF1 is to RIF1-dependent NHEJ in the context of CSR or if other key repair proteins are regulated by TLK1.

The STAT3 signaling pathway is important for immune system development and emerged as a clearly enriched pathway in our analysis of RNA-seq data. We demonstrated that LCLs are less responsive to IL6 stimulation, likely due to a reduction in IL6R levels (**Figure 3**). Whether this is a direct result of the TLK1 mutation or reflects the proband’s immunodeficient status, we cannot determine for certain. However, it is notable that a number of other signaling pathways are not perturbed, including the phosphorylation of two reported substrates, ASF1A and NEK1 (**Figure 2**), as well as PKC signaling that has been previously linked with neurodevelopmental disorders (**Supplementary Figure S5**)(55). Thus, we postulate that alterations in the STAT3 pathway could potentially contribute to the immune system phenotypes.

In a previous study of *TLK2* variants implicated in MRD57, we observed altered proximal interactions and partial mislocalization of TLK2 variant proteins. In contrast, we did not observe that with either *TLK1* variant, as both showed a very similar repertoire of proximal interactors compared to WT. This suggests potential differences in TLK1 and TLK2 regulation that remain to be further explored. Despite this, the proximal interaction profiles of TLK1 and TLK2 were highly similar, further supporting the proposition that they have a large degree of redundancy (**Figure 5**)(8). As *TLK2* mutations clearly impact neurodevelopment, we propose that the p.Q479E variant likely interferes with overall TLK1-TLK2 activity via homo and heterodimerization that impairs chromatin maintenance during brain development. Further analysis of the neurodevelopmental phenotypes of TLK1- and TLK2-deficient mice is therefore warranted.

In summary, our data strongly indicates that mutations in *TLK1* are likely to be relevant to rare NDDs and should be considered in clinical diagnostics. Further, TLK1, and potentially TLK2, may impact immune system development in certain contexts and additional work is needed to understand their cellular and developmental roles.

## Data availability statement

Data are available in public, open access repositories GEO (https://www.ncbi.nlm.nih.gov/geo/) ClinVar (https://www.ncbi.nlm.nih.gov/clinvar/) and PRIDE (https://www.ebi.ac.uk/pride/), included in the article or uploaded as supplementary information.

## Ethics approval

The study protocol was approved by the Mass General Brigham Institutional Review Board and informed consent was obtained from the participating family.

## Data Availability

Data are available in public, open access repositories GEO, ClinVar and PRIDE, included in the article or uploaded as supplemental material.

https://www.ncbi.nlm.nih.gov/geo/query/acc.cgi?acc=GSE241032

https://www.ncbi.nlm.nih.gov/clinvar/

https://www.ebi.ac.uk/pride/archive/projects/PXD019450

## Acknowledgements

We thank the Stracker lab and L. Deriano for helpful discussions, S. Khurana for antibody optimization, A. De Benedetti for the kind gift of p-NEK1 antibody, D. Zafra and J. Motley for collaboration agreement support, and the IRB Barcelona mass spectrometry core facility and the CCR Genomics facility for technical support. Thank you to the RGP and Broad Institute Center for Mendelian Genomics teams, particularly Melanie O’Leary and Heidi Rehm for project leadership, Ben Weisburd for tandem repeat analysis, Sarah Stenton for mitochondrial genome analysis, and Alba Sanchis-Juan, Harrison Brand, and Michael Talkowski for structural variant analysis.

## Supplemental Material

Supplementary materials and methods.

## Supplementary Figures

Figure S1: Uncropped western blots.

Figure S2: Uncropped western blots and gels from kinase assays and BioID.

Figure S3: Decipher analysis of TLK2.

Figure S4: BioID-western blot analysis and results comparisons.

Figure S5: Western blotting of PKC substrates.

## Supplementary Tables

Table S1: Primers used in this study.

Table S2: Antibodies used in this study.

Table S3: Differentially expressed genes and IPA analysis (excel).

Table S4: Analysis of gene specific SNPs in RNA-seq data.

Table S5: Seqr analysis of the *TLK1* mutation.

Table S6: BioID-MS data of TLK1 and NDD variants (excel).

