## Supplementary material for "Identification of a *de novo* mutation in *TLK1* associated with a neurodevelopmental disorder and immunodeficiency": Supplemental Material.pdf

Supplementary materials and methods.

**Supplementary Figures**

Figure S1: Uncropped western blots.

Figure S2: Uncropped western blots and gels from kinase assays and BioID.

Figure S3: Decipher analysis of TLK2.

Figure S4: BioID-western blot analysis and results comparisons.

Figure S5: Western blotting of PKC substrates.

**Supplementary Tables**

Table S1: Primers used in this study.

Table S2: Antibodies used in this study.

Table S3: Differentially expressed genes and IPA analysis (excel).

Table S4: Analysis of gene specific SNPs in RNA-seq data.

Table S5: Seqr analysis of the *TLK1* mutation.

Table S6: BioID-MS data of TLK1 and NDD variants (excel).

### Supplementary Materials and Methods

#### Genomic DNA Purification

Genomic DNA was purified from patient derived LCLs using the Gentra Puregene Cell kit (Qiagen) following manufacturer's instructions. Briefly, cells were washed and resuspended in Cell Lysis Solution. Next, proteins were removed by adding Protein Precipitation Solution and centrifuging at high speed. Clear supernatants were transferred to clean Eppendorf tubes containing isopropanol. After mixing gently by inversion, lysates were centrifuged, and the supernatant discarded. The DNA pellet was then washed with 70% ethanol, then air dried for 5 minutes. After this, DNA Hydration Solution was added and incubated at 65°C for 1 hour to dissolve the DNA. DNA was quantified using a DS-11 Spectrophotometer (DeNovix).

#### RNA isolation

Total RNA was extracted from LCLs using the SV Total RNA Isolation System (Promega) following manufacturer's instructions. Briefly, $2.5x10^6$ cells were lysed in 175 µL of β-mercaptoethanol-cointaining RNA Lysis Buffer and diluted by adding 350 µL of RNA Dilution Buffer. After mixing by inversion, lysates were placed in a heating block for 3 mins at 70°C. After centrifuging at 14,000 x g for 10 mins at RT, the cleared lysates were transferred to a fresh microcentrifuge tube and mixed with 200 µL of 95% ethanol. This mix was transferred to a spin column and centrifuged at 14,000 x g for 1 min. RNA was then washed in 600 µL of RNA Wash Solution, and again centrifuged at 14,000 x g for 1 min. After emptying the collection tube, 50 µL of DNAse Incubation Mix (40 µL Yellow Core Buffer, 5 µL 0.09 M $MnCl_2$, and 5 µL of DNase I enzyme) were placed directly to the membrane inside the spin basket and incubated for 15 mins at RT. After this incubation, 200 µL of DNase Stop Solution were added to the Spin Basket, and centrifuged at 14,000 × g for 1 min. Next, 600 µL of RNA Wash Solution were added and centrifuged at 14,000 × g for 1 min, followed by 250 µL of RNA Wash Solution and centrifuged at high speed for 2 mins. Next, RNA was eluted in 100 µL of Nuclease-Free Water. Lastly, RNA was precipitated in 0.1 M NaCl in 100% ethanol for 30 mins at –20°C, collected by centrifugation at 10,000 x g for 15 mins at 4°C, and resuspended in nuclease-free water. RNA quantification was done using a DS-11 Spectrophotometer (DeNovix).

**Isolation of DNA from agarose gels**

Briefly, bands were carefully excised with a clean scalpel, weighed and incubated at 50°C for 10 minutes with the corresponding amount of NTI Buffer. Next, DNA was bound at a silica membrane by placing it into a column provided by the kit. The membrane was then washed twice with NT3 buffer and dried by incubating at 70°C for 3 minutes. DNA was eluted in NE buffer and quantified using a DS-11 Spectrophotometer (DeNovix). PCR products were sequenced using a NextSeq (Illumina).

**RNA-seq analysis**

For gene expression analysis, contaminating adapter sequences were removed using Trimmomatic v0.36 and reads were mapped to the hg38 human reference genome using STAR v2.5.3 run in 2-pass mode(1,2). RSEM was then used for gene-level expression quantification, and data processing was performed in R statistical program environment(3). The edgeR program in R studio was used to determine differentially expressed genes between samples based on a false discovery rate (FDR) of 0.05 and a log FC of 1(4). Genes with a cpm (counts per million) value <0.5 in 3 samples were removed and the resulting counts data was normalized by trimmed mean of M-values (TMM) implemented in edgeR(5). Ingenuity Pathway Analysis (Qiagen) was run using DEGs (FDR<= 0.05). Data has been submitted to GEO (GSE241032).

Somatic mutations of selected genes were identified from RNAseq data using RNAmut(6). Briefly, for a list of genes of interest, the program uses FASTQ files from transcriptomic RNA-seq and creates an index file for the genes of interest and outputs detected mutations.

**Proximity-dependent biotin identification mass spectrometry (BioID-MS)**

AD-293 cells were seeded in 15 cm plates and transiently transfected the next day with 20 µg of BirA* plasmids using PEI and 150 mM NaCl as described(7). Medium was changed 6-8 hours post-transfection. 24 hours post-transfection, 50 µM of biotin (IBIAN Biotechnology) was added per plate. For mass spectrometry, 5x15 cm plates were used per condition. 48 hours post-transfection, the cells were harvested with Trypsin-EDTA (Sigma-Aldrich) and the 5 plates per condition were pooled together. Cell pellets were washed twice in cold PBS and lysed in 5 mL of cold lysis buffer (50 mM Tris-HCl pH 8.0, 150 mM NaCl, 0.1% SDS, 2 mM Mg2Cl, 1% Triton X-100 (Sigma-Aldrich), 1mM EDTA (Sigma-Aldrich), 1mM EGTA (SigmaAldrich), 1:2000 benzonase 25 U/mL (Sigma-Aldrich), 1x protease inhibitor cocktail (Roche) and 1x phosphatase inhibitor cocktails 2&3 (Sigma-Aldrich)). 100 µL of the lysate were retained for Western blotting analysis. The remaining lysate was incubated with streptavidin-sepharose beads (GE Healthcare 2-1206-010) during 3 hours in an end-over-end rotator at 4ºC in order to isolate the biotinylated proteins. The beads were washed once in lysis buffer and three times in 50 nM ammonium bicarbonate pH 8.3 buffer. Samples were snap-frozen and sent to the Mass Spectrometry & Proteomics Core Facility at IRB Barcelona for tryptic digestion and analysis. Tryptic digestion was performed directly on beads by incubation with 2 ug of trypsin in 50 mM NH4HCO3 at 37ºC overnight. The next morning, an additional 1 µg of trypsin was added and incubated for 2 h at 37°C. The digestion was stopped by adding formic acid to 1% final concentration. Samples were cleaned through C18 tips (polyLC C18 tips) and peptides were eluted with 80% acetonitrile, 1% TFA. Samples were diluted to 20% acetonitrile, 0.25% TFA, loaded into strong cation exchange columns (SCX) and peptides were eluted in 5% NH4OH, 30% methanol. Finally, samples were evaporated to dry, reconstituted in 50 µL and diluted 1:8 with 3% acetonitrile, 1% formic acid aqueous solution for nanoLC-MS/MS analysis. The nano-LC-MS/MS was set up as follows. Digested peptides were diluted in 3% ACN/1% FA. Sample was loaded to a 300 µm × 5 mm PepMap100, 5 µm, 100 Å, C18 µ-precolumn (Thermo Scientific) at a flow rate of 15 µl/min using a Thermo Scientific Dionex Ultimate 3000 chromatographic system (Thermo Scientific). Peptides were separated using a C18 analytical column Acclaim PEPMAP 100 75 µm x50 cm nanoviper C18 3 µm 100A (Thermo Scientific) with a 90 min run, comprising three consecutive steps with linear gradients from 3 to 35% B in 60 min, from 35 to 50% B in 5 min, and from 50% to 85% B in 2 min, followed by isocratic elution at 85% B in 5 min and stabilization to initial conditions (A= 0.1% FA in water, B= 0.1% FA in CH3CN). The column outlet was directly connected to an Advion TriVersa NanoMate (Advion) fitted on an Orbitrap Fusion Lumos™ Tribrid (Thermo Scientific). The mass spectrometer was operated in a data-dependent acquisition (DDA) mode. Survey MS scans were acquired in the Orbitrap with the resolution (defined at 200 m/z) set to 120,000. The lock mass was user-defined at 445.12 m/z in each Orbitrap scan. The top speed (most intense) ions per scan were fragmented by CID and detected in the linear ion trap. The ion count target value was 400,000 and 10,000 for the survey scan and for the MS/MS scan respectively. Target ions already selected for MS/MS were dynamically excluded for 15s. Spray voltage in the NanoMate source was set to 1.60 kV. RF Lens were tuned to 30%. Minimal signal required to trigger MS to MS/MS switch was set to 5,000. The spectrometer was working in positive polarity mode and singly charge state precursors were rejected for fragmentation. We performed a twin database search with two different softwares, Thermo Proteome Discoverer v2.3.0.480 (PD) and MaxQuant v1.6.6.0 (MQ). The search engine nodes used were Sequest HT for PD and Andromeda for MQ. The databases used in the search was SwissProt Human (release 2019 01) including contaminants and TLK1 and TLK2 proteins. We run the search against targeted and decoy databases to determine the false discovery rate (FDR). Search parameters included trypsin enzyme specificity, allowing for two missed cleavage sites, oxidation in M and acetylation in protein N-terminus as dynamic modifications. Peptide mass tolerance was 10 ppm and the MS/MS tolerance was 0.6 Da. Peptide were filtered at a false discovery rate (FDR) of 1 % based on the number of hits against the reversed sequence database. For the quantitative analysis, contaminant identifications were removed and unique peptides (peptides that are not shared between different protein groups) were used for the quantitative analysis with SAINTexpress-spc v3.6.1(8). SAINTexpress compares the prey control spectral counts with the prey test spectral counts for all available replicates. For each available bait and for each available replicate, we took as prey count the maximum count result between PD and MQ. Once obtained this combined dataset, we ran the SAINTexpress algorithm with TLK1 samples and a number of controls samples from previous experiments in the same cell type (n=45 total). High confidence interactors were defined as those with a SAINT score of 0.7 or greater. Raw data is available in the PRIDE repository, accession number PXD019450.

**Western blotting**

For affinity purification (AP), 40 µg of input protein and 20 µL of Strep-AP elution, with 6X SDS (0.2% bromophenol blue and β-mercaptoethanol), were separated by SDS-PAGE and transferred to nitrocellulose membranes (0.2 or 0.45 µm pore, Amersham Protran; Sigma-Aldrich). For detection of streptavidin, polyvinylidene fluoride membranes (0.45 µm pore, Immobilon-P, Merck) were used. Membranes were blocked and antibodies prepared in 5% non-fat milk in PBS-Tween20 (PBS-T). Primary antibodies were detected with the appropriate secondary antibodies conjugated to horseradish peroxidase (HRP) and visualized by ECL-Plus (GE Healthcare).

For LCL western blots, cells were lysed for 20-30 min in RIPA buffer (150 mM NaCl, 50 mL Tris pH 7.4, 1% NP-40, 0.5% DOC, 0.1% SDS, 1X protease and phosphatase inhibitors) and briefly sonicated. 10-40 µg of protein were separated by SDS-PAGE using Mini-PROTEAN TGX Gels (Bio-Rad) and transferred to PVDF (0.45 µm pore, Bio-Rad) or Nitrocellulose (0.2 µm pore, GE Healthcare Life Sciences) membranes using the Trans-Blot Turbo transfer system (Bio-Rad). Membranes were blocked and antibodies prepared in EveryBlot Blocking Buffer (Bio-Rad).

Primary antibodies were detected with the appropriate secondary antibodies (IRDye 800CW Goat anti-rabbit IgG, IRDye 800CW Goat anti-mouse IgG, IRDye 680RD Goat anti-rabbit IgG, or IRDye 680RD Goat anti-mouse IgG (LI-COR)) and visualized using an Odyssey CLx (LI-COR). Antibodies used are provided in **online supplementary table S2.**

***In vitro* Kinase assays**

*In vitro* kinase assays were performed as previously described(7). After Strep-AP of pcDNA3.1 N-SF-TAP TLK1 from AD-293 cells, 200 µg of Strep-AP were incubated with 2 µCi 32P-γ-ATP, 100 µM cold ATP, 1 µg of purified ASF1A protein in 12 µL of kinase buffer (50mM Tris-HCl pH 7.5, 10 mM MgCl2, 2mM DTT, 1X protein inhibitor cocktail (Roche) and 1X phosphatase inhibitor cocktails 2&3 (Sigma-Aldrich)). The reaction was incubated at 30ºC for 30 min. After that, the reaction was stopped by adding 4 µL of Sample Buffer (6X SDS, 0.2% bromophenol blue and β-mercaptoethanol), and boiled for 5-10 mins at 95ºC. Samples were analyzed on SDS-PAGE, stained with Coomassie Blue for 1 hour, washed 4 times with destaining buffer (10% acetic acid, 40% methanol, and 50% $H_2O$) and vacuum dried with an SGD2000 (Savant) for 2 hours at 60ºC. TLK1 and ASF1A phosphorylation were measured using a Typhoon 8600 Variable Mode Imager (Molecular Dynamics) and band intensity quantified using ImageJ(9).

**Comet assay**

Comet assays were performed according to manufacturer’s instructions (Trevigen). Briefly, cells were mixed with low melting agarose, and allowed to solidify onto comet slides for 45 minutes. For Alkaline Comet assay, cells were incubated in Lysis Solution for 1 hour at 4°C, and then immersed in freshly prepared Alkaline Unwinding Solution, pH > 13 (200 mM NaOH, 1 mM EDTA) for 20 minutes at room temperature. The slides were then placed in a CometAssay ES II electrophoresis system (Biotechne), immersed in cold Alkaline Electrophoresis Solution, pH > 13 (200 mM NaOH, 1 mM EDTA), and a voltage of 21 V was applied for 1 hour. Next, the slides were gently immersed twice in $dH_2O$ for 5 minutes each, and then in 70% ethanol for another 5 minutes and dried overnight at room temperature. Finally, slides were stained with 1X SYBR Gold

(Invitrogen) for 30 minutes at room temperature and briefly rinsed in $dH_2O$. To assess DNA damage induced by γ-ray irradiation and repair capability, cells were treated with 2-Gy γ-rays (XRAD 320, Precision X-ray, Inc) on ice to prevent DNA damage repair. Residual DNA damage was evaluated after 15- and 30-minutes incubation at 37°C. Images were obtained on an AXIO Imager.Z2 (Zeiss) using a 10X objective. At least 50 cells were counted in each condition over two independent experiments. Experiments were quantified using CometScore 2.0 software (TriTek Corp.). Statistical analysis was performed using two-tailed unpaired Student's t test with Welch's Correction.

**Proliferation assay**

Proliferation was assessed using the Click-iT Plus EdU Flow Cytometry Assay Kit (Invitrogen) following manufacturer's instructions. Briefly, cells were labeled with 10 μM EdU for 1 hour at 37°C. Next, cells were washed in 1% BSA in PBS = and cell pellets were dislodged in 100 μL of Click-iT fixative (containing 4% PFA). Cells were incubated for 15 minutes at room temperature, protected from light. Cells were washed in 1% BSA in PBS and resuspended in 100 μL of 1X Click-iT saporin-based permeabilization and wash reagent. Cells were incubated for 15 minutes on ice. Next, we performed the Detect ClickiT EdU reaction for 30 minutes at room temperature, protected from light. Cells were washed once in 1X Click-iT permeabilization and wash reagent and stained with DAPI (0.5 μg/mL) in 1% BSA, 0.02% NaAz in PBS for 30 minutes at 37 ºC. Samples were acquired in a CytoFLEX LX (Beckman Coulter) flow cytometer analyzer.

**Cloning and site-directed mutagenesis**

The human *TLK1* variant 5 cDNA was obtained in a Gateway-compatible pENTR223 vector (Sigma-Aldrich Mission cDNA library) and introduced into the N-SF-TAP-DEST vector (a kind gift from J Gloeckner)(10) by recombination reaction (300 ng TLK1-pENTR223, 300 ng N-SF-TAP-DEST, 4 μL Gateway buffer (Invitrogen, Carlsbad, CA, USA) and 4 μL clonase LR (Invitrogen)) and incubated at 25 °C for 1 h. Proteinase K (2 μL) (Sigma-Aldrich) was added and incubated at 37 °C for 10 min to stop the reaction. Competent *Escherichia coli* DH5α (Sigma-Aldrich) were transformed with the resulting recombination reaction and selected with kanamycin. The TLK2 kinase dead mutation (D592V) was generated as previously described(11). Plasmids were checked by restriction digests and sequencing (Macrogen) and DNA prepared with MaxiPreps (Promega, Madison, WI, USA).

To generate BioID constructs, *TLK1* was amplified with a forward primer containing 5′ AscI and a reverse primer containing 3′ NotI restriction sites using KOD Hot Start DNA Polymerase (Millipore) and cycling conditions recommended from the manufacturer (polymerase activation at 95 °C for 2 min, denaturation at 95 °C for 20 s, annealing at 55 °C for 10 s and extension at 70 °C for 50 s, repeated for 40 cycles). PCR products were purified using the PureLink Quick Gel Extraction Kit (Invitrogen) and cloned into pCR2.1-TOPO vector (Invitrogen). Top 10 competent *E. coli* cells (Invitrogen) were transformed with pCR2.1-TLK1 and colonies were selected in carbenicillin. TLK1 was cut from the pCR2.1-TOPO vector by restriction digest with AscI (New England BioLabs (NEB), Ipswich, MA, USA) and NotI-HF (NEB), purified using the PureLink Quick Gel Extraction Kit (Invitrogen) and ligated into pcDNA5/FRT/TO-N-FLAG-hBirA* using a Quick Ligation Kit (NEB). Top 10 competent *E. coli* cells (Invitrogen) were transformed and carbenicillin selected. The constructs were confirmed by restriction digestion with *Asc*I (NEB) and *Not*I-HF (NEB) and sequencing (Macrogen).

*TLK1* mutations were generated using the QuickChange Lightning site-directed mutagenesis kit (Agilent Technologies) on the plasmids N-SF-TAP-DEST-TLK1-WT and pcDNA5/FRT/TO-N-FLAG-hBirA*-TLK1-WT following manufacturer’s instructions. The primers used are provided in **online supplementary table S1**. Constructs were subsequently transfected in AD-293 cells as previously described(7).

Figure 2C

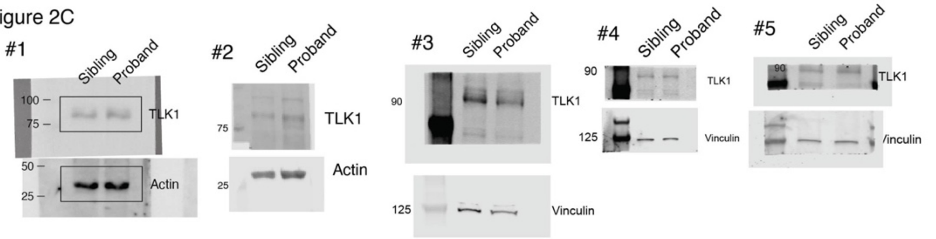

Figure 2D

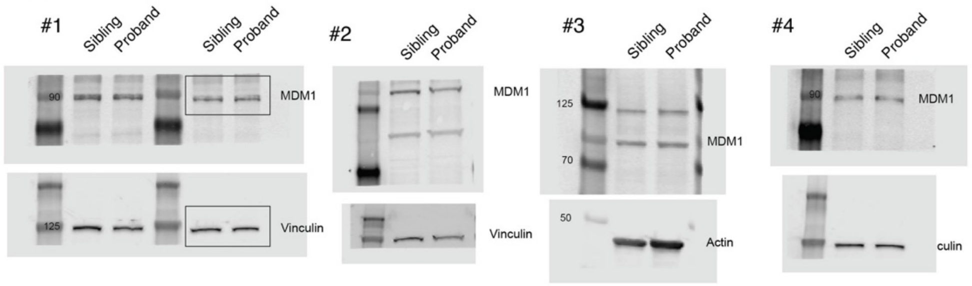

Figure 2E

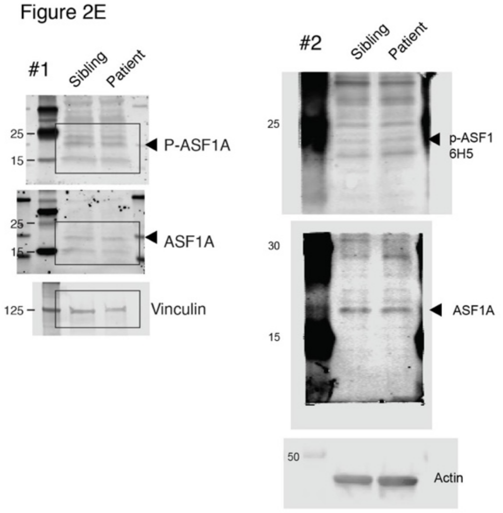

Figure 2F

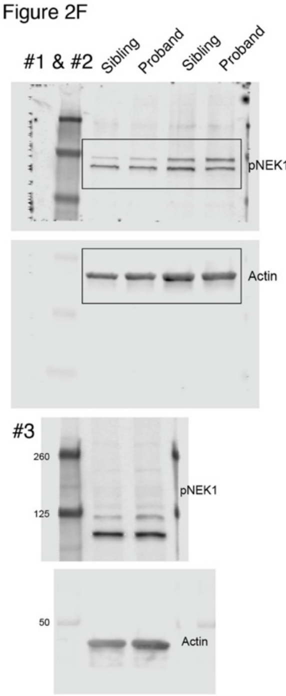

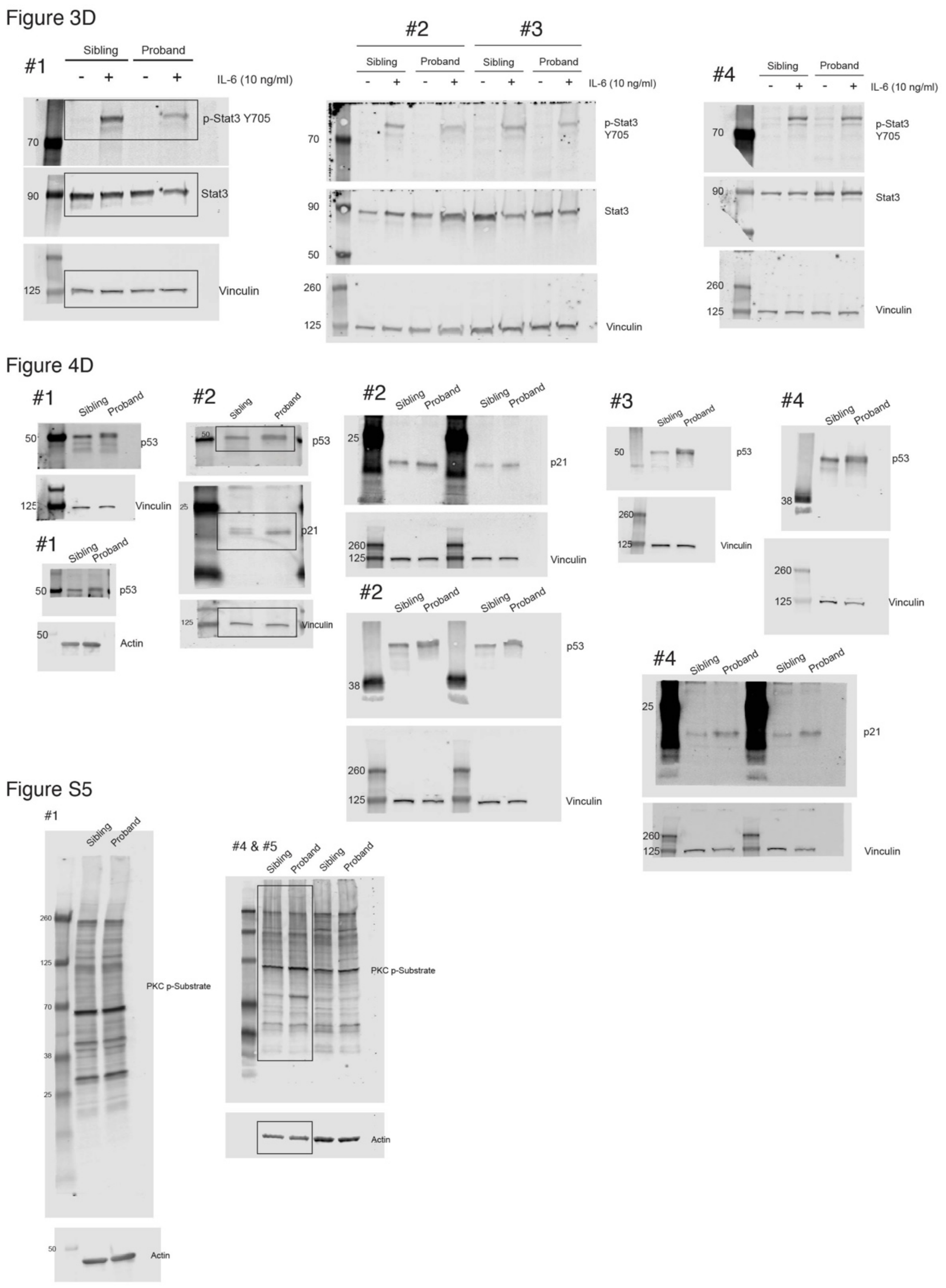

**Figure S1: Uncropped Western blots.** Figure panels where data was used are indicated. Replicates used for quantification are included and regions cropped for figures are shown.

Figure 5B

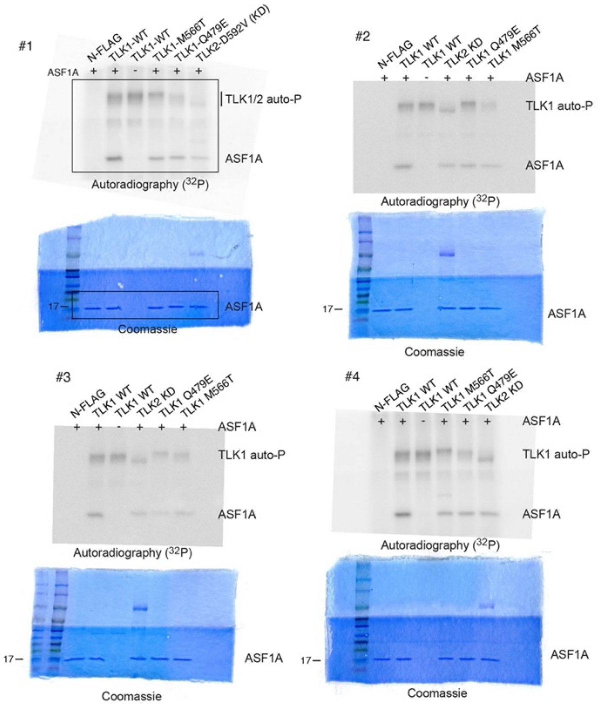

Figure 5D

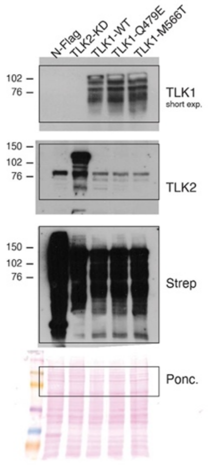

Figure S4A

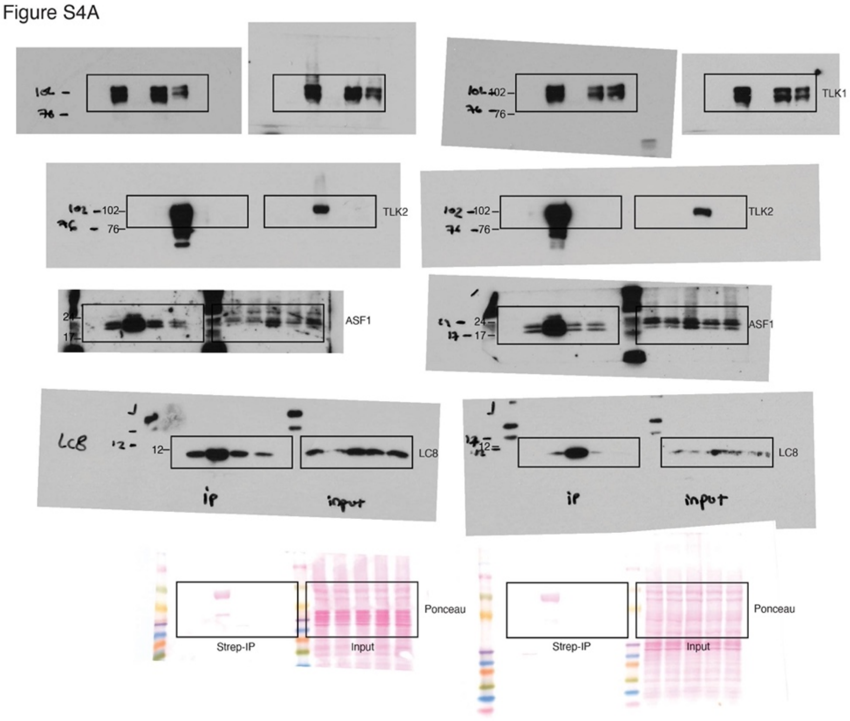

**Figure S2: Uncropped Western blots, autoradiographs and gels from kinase assays and BioID.** Figure panels where data was used are indicated and region cropped for figures is shown.

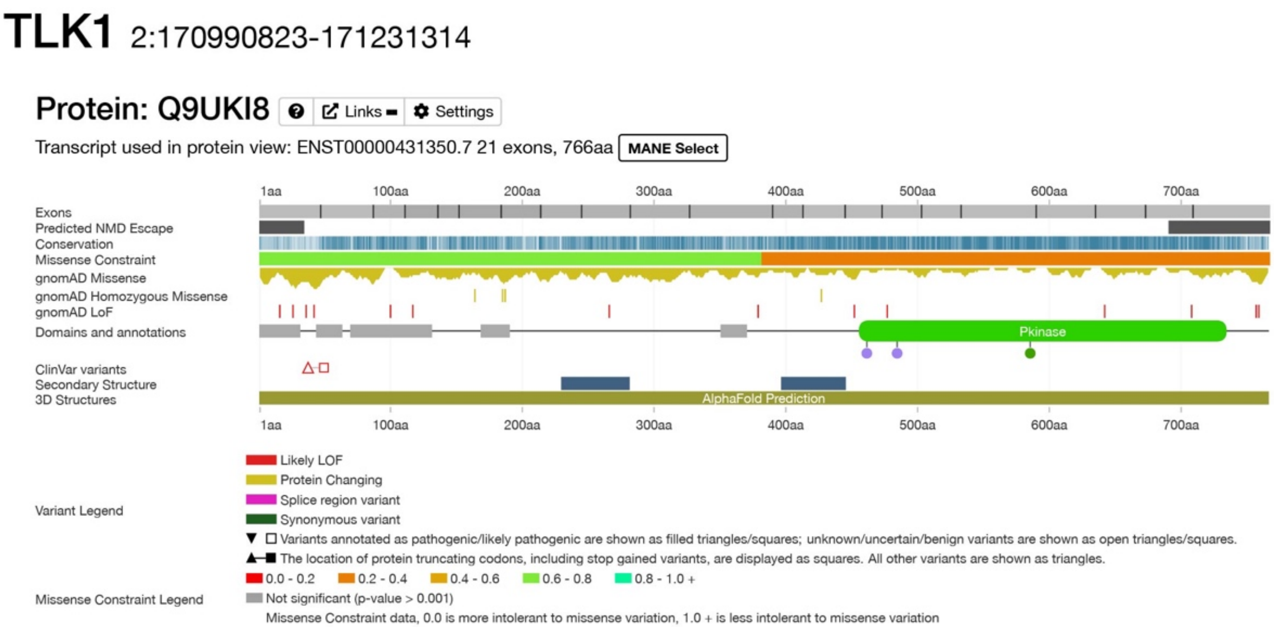

**Figure S3: Decipher analysis of TLK1.** Note the high level of missense constraint in the kinase domain (www.deciphergenomics.org).

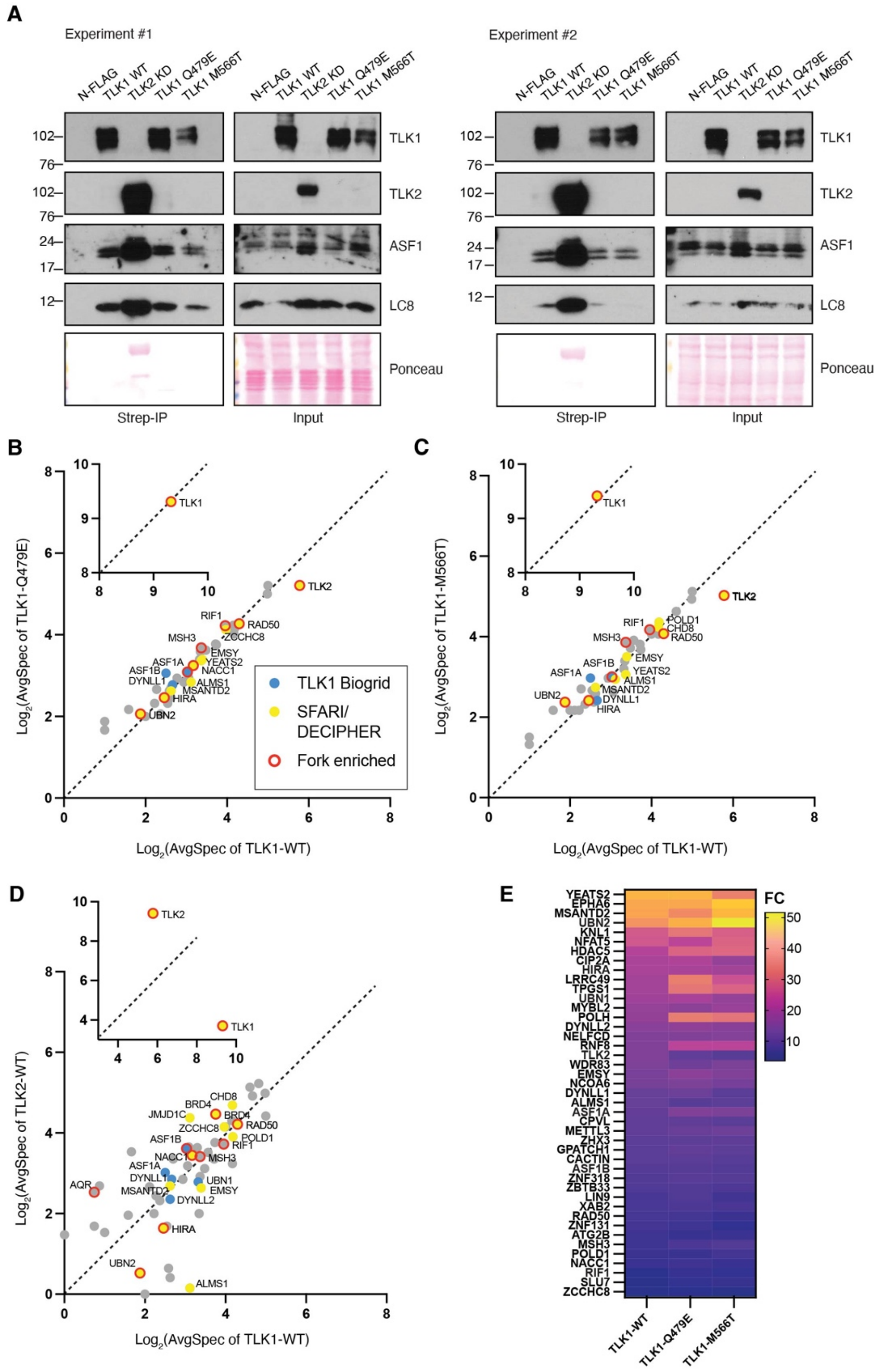

**Figure S4: BioID-western blot analysis and results comparisons. A.** Western blotting of BioID inputs and eluates for the indicated bait proteins and interactors. **B.** Scatter plot comparing BioID hits from TLK1-WT and TLK1-Q479E that scored with a BFDR of < or = to 0.06 in at least one of the samples. Proteins reported to interact with TLK1 (www.Biogrid.com), that are present in the SFARI or Decipher databases or are known to be enriched at replication forks(12) are indicated. **C.** Same as B for TLK1-M566T. **D.** Same as B but comparing TLK1-WT to TLK2-WT based on previous data(7). **E.** Heatmap comparing the fold change (FC) of hits compared to controls.

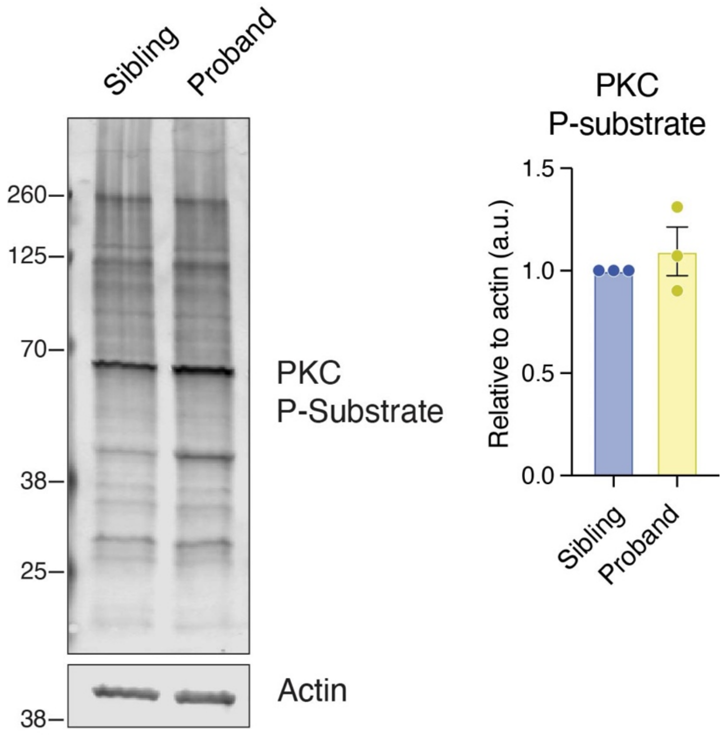

**Figure S5:** Western blotting of PKC substrates from lysates of the patient-derived LCLs. Quantification of lanes from N=3 blots from biological replicates are shown on the right. All uncropped blots are shown in Figure S1.

| **Target gene** | | **Sequence 5' to 3'** | **Product size** |
|---|---|---|---|
| TLK1-Q479E | Fwd | aggcttttgacctttatgaagaaagatatgctgctgtgaag | N/A |
| | Rev | cttcacagcagcatatctttcttcataaaggtcaaaagcct | |
| TLK1-M566T | Fwd | gaaagaagctcggtctattgtaacgcagattgtaaatgcac | N/A |
| | Rev | gtgcatttacaatctgcgttacaatagaccgagcttctttc | |
| TLK1 seq | Fwd | CACATACGTGAGCTGAAAAG | N/A |
| TLK2 KD seq | Fwd | CTTTTCACTGGATACTGAC | N/A |
| *TLK1* | Fw | ACCATGGAATTTCTGTCTTCTGA | 411 |
| | Rv | GGGTGATCCAGTTCTTTGTGTA | |

| | | | |
|---|---|---|---|
| *MDM1* | Fw | GCCAGTTTAGGGCTAGGTATG | 512 |
| | Rv | TGTCGTCCTCCTCCTCTTT | |
| *PSMB8* | Fw | CCTCACCCTCATACTCCCTAAA | 404 |
| | Rv | GTAAGGCACCTGGAAGAAGATG | |

**Table S1: Primers used for mutagenesis, sequencing or gDNA amplification.** Fw, forward; Rv, reverse.

| Antigen | Species | Source & reference | Dilution |
|---|---|---|---|
| TLK1 | Rabbit | Cell Signaling #4125 | 1:1000 (WB) |
| TLK2 | Rabbit | Bethyl Laboratories A301-257A | 1:1000 (WB) |
| ASF1 | Rabbit | Cell Signaling #2990 | 1:1000 (WB) |
| ASF1-pS166 (1E6 and 6H5) | Mouse | Genscript | 1:500 (WB) |
| Actin | Mouse | Sigma-Aldrich A4700 | 1:1000 (WB) |
| Actin | Mouse | Millipore Sigma MAB1501, clone C4 | 1:3000 (WB) |
| GAPDH | Mouse | Santa Cruz sc-47724, clone 0411 | 1:1000 (WB) |
| FLAG | Mouse | Sigma-Aldrich F3165, clone M2 | 1:5000 (WB) |
| Strep-tag | Mouse | IBA GmbH 2-1509-001 | 1:1000 (WB) |
| MDM1 | Rabbit | Sigma HPA041594 | 1:1000 (WB) |
| P53 | Mouse | Cell Signaling #48818 | 1:1000 (WB) |
| P21 | Mouse | Santa Cruz sc-53870 | 1:500 (WB) |
| P-STAT3 | Rabbit | Cell Signaling #9145 | 1:2000 (WB) |
| STAT3 | Mouse | Cell Signaling #9139 | 1:1000 (WB) |
| pPKC Substrate | Rabbit | Cell Signaling #6967 | 1:1000 (WB) |
| Vinculin | Rabbit | Cell Signaling #13901 | 1:2000 (WB) |
| NEK1-pT141 | Rabbit | Kind gift from Arrigo de Benedetti | 1:1000 (WB) |
| LC8 | Rabbit | Abcam ab51603, clone EP1660Y | 1:1000 (WB) |
| Protein A/G anti-rabbit HRP | | Thermo Fisher Scientific 32490 | 1:15000 (WB Nitro)<br>1:30000 (WB PVDF) |
| goat anti-mouse IgG HRP | | Thermo Fisher Scientific 31430 | 1:15000 (WB Nitro)<br>1:30000 (WB PVDF) |
| IRDye 800CW Goat anti-Rabbit IgG | | LI-COR Biosciences 926-32211 | 1:10000 (WB PVDF) |
| IRDye 800CW Goat anti-Mouse IgG | | LI-COR Biosciences 926-32210 | 1:10000 (WB PVDF) |
| IRDye 680RD Goat anti-Rabbit IgG | | LI-COR Biosciences 926-68071 | 1:10000 (WB PVDF) |
| IRDye 680RD Goat anti-Mouse IgG | | LI-COR Biosciences 926-68070 | 1:10000 (WB PVDF) |

WB, western blot; IF, immunofluorescence ; FC, flow cytometry

**Table S2: Antibodies used in this study.**

**Table S3: Differentially expressed genes and IPA analysis (Excel).**

| RNAseq ID | Gene | Mutation | ProtMut | MutReads | WTReads | VAF | Info |
|---|---|---|---|---|---|---|---|
| P007-S15 | *TLK1* | chr2:171007045-171007045_Sub:G>C | Q479E | 67 | 61 | 0.523 | Neutral |
| P007-S16 | *TLK1* | chr2:171007045-171007045_Sub:G>C | Q479E | 80 | 48 | 0.625 | Neutral |

**Table S4: relative expression of TLK1 and MDM1 variants.** The number of reads for the mutant (Mut) and WT TLK1 from RNAseq of LCLs derived from the proband are shown. No reads for the variant *MDM1* allele were detected in samples from the unaffected sibling.

| **Predictor** | **Score** |
|---|---|
| Cadd | 23 |
| Revel | 0.38 |
| Primate Ai | 0.88 |
| Mpc | 1.6 |
| Splice Ai | 0 |
| Eigen | 3.2 |
| Polyphen | Benign |
| Sift | Damaging |
| Mut Taster | Disease causing |
| Fathmm | Tolerated |
| Metasvm | Tolerated |
| Gerp Rs | 4.95 |
| Phastcons 100 Vert | 1 |

**Table S5: *In silico* predictions of the TLK1 Q479E impact using seqr.** The predictor, related score and color-coded impact are shown. Green indicates tolerated, Red indicates damaging and Yellow indicates potentially damaging.

**Table S6: BioID-MS data of TLK1 and NDD variants (Excel).**
